# An evaluation of combined objective neurophysiologic markers to aid assessment of prolonged disorders of consciousness (PDoC)

**DOI:** 10.1101/2024.10.09.24315104

**Authors:** N. du Bois, J. Hill, A. Korik, D. Hoad, L. Bradley, S. Judge, T. M. Vaughan, J. R. Wolpaw, D. Coyle

## Abstract

**Objective:** Clinical assessments of individuals with Cognitive-Motor Dissociation (CMD) following brain injury are challenging and prone to errors. This prompts investigation of objective, movement-independent neurophysiological markers using electroencephalography (EEG)-based Brain-Computer Interface (BCI) technology. The current pilot study involving adults with prolonged disorders of consciousness (PDoC) investigated the combination of Motor-Imagery BCI (MI-BCI) training and auditory evoked Event Related Potentials (ERPs) using an oddball paradigm to produce complementary biomarkers to improve evaluation of awareness in PDoC.

**Approach:** EEG data (16 channels) were collected from participants with Unresponsive Wakefulness Syndrome (UWS, *n* = 2), Minimally Conscious State (MCS, *n* = 3), and Locked-In Syndrome (LIS, *n* = 4). The MI-BCI involved assessing sensorimotor rhythm modulation, motor-imagery training with and without auditory feedback, and motor-imagery responses to closed questions over 12 sessions each lasting ∼1hour. The oddball protocol was also deployed in 2-3 of those sessions, with ∼10 days between first and last, featuring auditory stimuli, comprising two 5-minute sets of standard, deviant beeps plus novel sounds, in a structured ratio. We expected those with the lowest levels of awareness would have reduced ERP components, with highest latencies to peak, as well as lowest accuracy in the motor imagery BCI protocol – and that trends across these metrics would be observed across the three patient groups based on their clinical diagnoses.

**Main results:** Significant differences in mean N1 component latencies and mean MI Decoding Accuracies (DA, for significant runs) occurred between groups – with shorter N1 latencies for the LIS and MCS groups than for the UWS group (LIS vs. UWS and MCS vs. UWS, *p* < 0.001), and higher DA for the LIS group compared to MCS and UWS (*p* < 0.001). Mean DA were found to have a significant negative correlation with mean N1 latencies (two-tailed, *p* = 0.017).

**Significance:** The results indicate that neurophysiological markers from the concomitant application of an MI-BCI and auditory-oddball paradigm can augment standard clinical assessments by providing objective measures that produce robust evidence of awareness in people with PDoC.

## 1 Introduction

In the context of medical practice, Disorders of Consciousness (DoC) describes a continuum of alterations of consciousness that occur following acquired brain injury. When a DOC persists beyond four-weeks post-injury, the condition is then described as a Prolonged Disorder of Consciousness (PDoC) [1]. A DoC has two components: wakefulness and awareness. Wakefulness is associated with the brain’s arousal systems, primarily driven by subcortical structures (broadly the brainstem and thalamus) – while, awareness relies on a distributed frontoparietal network for the necessary complex processing in higher-cortical areas, related to the perception of, and interaction between, the self and the environment [2–4]. The state or quality of wakefulness or awareness of an individual with a DoC can range from complete unconsciousness, where both wakefulness and awareness are absent, as in coma, which is usually acute and short-term – to a state of wakefulness with absent awareness, as in Unresponsive Wakefulness Syndrome (UWS), or to a state of wakefulness with minimal or fluctuating awareness, as in the Minimally Conscious State (MCS) [1]. Individuals diagnosed with UWS have a loss of functional connectivity in the long-range cortico-cortical and cortico-thalamo tracts, while individuals diagnosed with MCS have preserved functional connectivity in the frontoparietal network (FPN). The emergence from UWS to MCS depends on restored functional connectivity in the FPN [4]. The MCS diagnosis extends along a continuum of cognitive function, where MCS minus (MCS-) and MCS plus (MCS+) correspond to the integrity of the FPN [5,6]. Separate to a DoC, Locked-In Syndrome (LIS) [7,8], describes a neurological condition that occurs following an acquired brain injury that causes severe damage to the brainstem while leaving other brain areas intact, for example a brainstem infarct (i.e., brainstem stroke), resulting in quadriplegia and anarthria – leaving the individual with fully preserved consciousness but unable to move (apart from eye-movements).

Despite recent research demonstrating an ability to detect consciousness using non-invasive neuroimaging techniques, and protocols that do not require a behavioural response [9], the most commonly used approaches to determine states of DoC and LIS are the Coma Recovery Scale – Revised (CRS-R) [10], or the Wessex Head Injury Matrix (WHIM) [11], which are standardised assessment tools that evaluate the neurobehavioral functioning of patients with DoC. Such tools aim to differentiate between DoC states by assessing various domains of functioning, including auditory, visual, motor, oromotor, communication, and arousal levels. The American Academy of Neurology (AAN), the European Academy of Neurology (EAN) and the UK Royal College of Physicians (RCP) disagree on the use cases of neuroimaging tools to aid in the diagnosis of DOC. While the AAN and EAN express interest in the potential additive value of neuroimaging techniques (under varying conditions), concerns common to all three authorities include cost-effectiveness, practical implementation, and more robust evidence [9]. Owen et al. (2006) [12], pioneered the use of neuroimaging techniques to assess for evidence of consciousness in people diagnosed with UWS. This seminal functional magnetic resonance imaging (fMRI) case-study demonstrated the ability of a patient diagnosed with UWS to covertly follow commands, using an imagined movement protocol involving two tasks; imagining (1) playing tennis and (2) walking around their room – providing the first indication that a patient diagnosed with a PDoC assessment of UWS could intentionally modulate brain activity. This foundational work was advanced by Monti et al. (2010) [13], who employed similar fMRI techniques to investigate communication in PDoC patients through yes/no biographical questions, with some patients successfully modulating their brain activity to respond. The first evidence to support the efficacy of electroencephalographic (EEG) technology to measure the intentional modulation of sensorimotor rhythms (SMR) during imagined movement (i.e., motor-imagery), in a cohort of three UWS patients, was provided by Cruse et al. (2011) [14]. The application of EEG-based motor-imagery (MI) protocols lowers the cost and increases accessibility, as compared to fMRI, of potential technology that might add diagnostic value in an assessment of a patient with a DoC – addressing the concerns of the AAN, EAN, and RCP, relating to cost-effectiveness and practical implementation.

Subsequent studies have reinforced the potential of EEG-based MI protocols to detect awareness in patients with disorders of consciousness [15–17]. Relevant to the current pilot, Coyle et al. (2015) [18] demonstrated that training patients with a diagnosis of MCS (*N* = 4) to improve their sensorimotor modulation using an EEG-based MI-BCI with auditory feedback is feasible – and could potentially serve as a communication tool for this patient cohort. Building on this research, the MI-BCI protocol was further developed to include a question and answer (Q&A) task, and research evaluating the efficacy of the full protocol is currently being conducted with a large cohort of PDoC and LIS patients, across fifteen UK sites [19,20]. The full MI-BCI protocol [20] provides three possible imagined movement combinations for a two-class movement classification: left vs. right arm, right arm vs. feet, and left arm vs. feet. Each participant uses a specific combination throughout their sessions.

While recent advances in wearable EEG-based BCIs (eBCIs) stand to further address issues of cost-effectiveness and accessibility, and therefore, accelerate the adoption of validated BCI protocols in healthcare in the near future [21,22] – to allay concerns related to the integrity of the evidence for the additive value of neuroimaging protocols to aid a diagnosis of DoC or LIS, further research is required. Specifically, to validate and establish the reliability and utility of these protocols will require the development of a battery of EEG paradigms and measures to assess different aspects of neural function, with demonstrable internal consistency. Kim et al. (2022) [23], has recently progressed this approach using a protocol that comprises both passive (auditory oddball) and active (covert command following) EEG paradigms, to investigate neurophysiological markers that reflect differences in the cognitive state of children with brain injury (aged 8-18 years: brain injured, *n* = 31; control, *n* = 13). Key EEG markers identified using this protocol were the N1 and the P3 components of the stimulus evoked event-related potential (ERP) — the former reflects early sensory stimulus processing and the latter a higher-cognitive function that indicates the orientation of attention to a salient stimulus. Both markers were found to increase in magnitude as cognitive recovery progressed, with the N1 demonstrating greater separation of participant groups, while the P3 magnitude provided a more reliable indicator of an individual’s ability to follow commands. The Kim et al. (2022) [23] study represents the largest cognitive ERP research to date focusing on paediatric brain injury and provides objective, movement-independent EEG markers to aid in the evaluation of cognitive state – offering robust evidence supporting the use of neuroimaging tools in the process of diagnosing disorders of consciousness (DoC), in a paediatric cohort with brain injury.

Building on these research advances, the current pilot study, conducted with a cohort of patients (*N* = 9) recruited to participate in the ongoing large-scale EEG-based MI-BCI study (Clinical Trials Registration number: NCT03827187), employed the passive auditory oddball protocol (demonstrated in a paediatric population, by Kim et al. (2022) [23]), alongside the MI-BCI protocol [19,20,24] in the same individuals. There were three aims of this study: first, to determine whether the ERP markers identified by Kim and colleagues can differentiate between PDoC patients with a diagnosis of either UWS or MCS, as well as those with LIS; and second, to establish whether there is a consensus between these potential ERP markers and motor-imagery ability, within an adult cohort of PDoC and LIS patients and thirdly, to determine whether this combined auditory oddball and MI-BCI protocol can exploit the same EEG markers identified by Kim et al. (2022) [23] to distinguish between adult PDoC (UWS, MCS) and LIS patients. Demonstrating correlation between these EEG-markers and motor imagery ability by evaluating the internal consistency of these neurophysiological markers, within an adult cohort of PDoC and LIS patients, will help establish robust evidence for the use-case of neuroimaging tools to aid a diagnosis of DoC – addressing key concerns raised by the AAN, EAN and RCP [9].

## 2 Methods

### 2.1 Participants

There were nine participants in this pilot study (UWS (*n* = 2), MCS (*n* = 3), and LIS (*n* = 4)), recruited to participate in an ongoing large-scale study (Clinical Trials Registration number: NCT03827187). Participant details are presented in Supplementary Table 2. The study was ethically approved by the Health Research Authority (HRA) and Health and Care Research Wales (HCRW) Research Ethics Committee (REC), of the National Health Service (NHS), and was conducted in accordance with the Declaration of Helsinki. The principal investigators (PIs) at the participating NHS sites are Neurology and Neurorehabilitation consultants in charge of PDoC patients’ care. The PIs, or a member of the care team, made the initial decision about which patients were recruited for the study. Consent to participate was given where possible by the patient, or proxy consent was provided by the family, and the PIs informed the research team of the patient’s current PDoC diagnosis, i.e., whether they were in a UWS, MCS, or LIS state. Participants were visited as inpatients at participating NHS hospitals, or in care homes or patients’ own homes in the United Kingdom (UK).

### 2.2 Experiment protocol and stimuli

The EEG auditory oddball paradigm involves a passive listening task that presents a rapid randomised sequence of auditory stimuli – including both standard and deviant stimuli, with the latter presented less frequently than the former. The paradigm is used to trigger auditory evoked ERPs that demonstrate robust deviant versus standard ERP differences [23,25]. Here the protocol comprised 2-3 sessions, with 10 (+/- 3) days between the first and last session. Each session comprised a two-minute baseline resting state recording, followed by two 5-minute oddball sets of standard and deviant beeps plus novel sounds, delivered in a structured standard-deviant-novel ratio of 27:8:6. The oddball task and stimuli employed, are described in Kim et al. (2022) [23]. The protocol contained five different sets of 1-minute 22s in duration, each a different sequence of standard-deviant-novel stimuli, and each with different novel sounds – but all having the same ratio of standards, deviants and novel stimuli, presented with an inter-stimulus interval of 1-second. Sets 1 and 2, 3 and 4, 1 and 5, were selected for the first session, second session and third session respectively (not all participants completed the third session). The selected set was looped for a 5-minute interval.

#### Auditory oddball stimuli

The stimuli were square-wave beeps with a duration of 340ms. The fundamental frequency for the standard stimuli was 400Hz and for the deviant stimuli was 575Hz. The novel stimuli were sounds like a doorbell, a horn, a cat meowing etc., and were designed with the aim to evoke a strong P3a component (reflective of stimulus-driven frontal attention mechanisms during task processing [26]).

#### Motor-imagery Brain-Computer Interface (MI-BCI) study outline

Two of three potential imagined movement combinations were used for a two-class movement classification: left-vs. right-arm, right-arm vs. feet, and left-arm vs. feet. The study had three phases. Phase I (sessions 1-2) assessed participants’ ability to modulate brain activity to achieve significant decoding accuracy (DA), requiring a peak DA during the task period significantly higher than both the baseline period and randomly permutated samples. Phase II (sessions 3-6) involved MI-BCI training with neurofeedback to train participants to modulate their brain activity. Phase III (sessions 7-10) evaluated participants’ MI-BCI responses to closed questions, categorised as biographical, numerical, logical, and situational awareness [19,20].

### 2.3 EEG data acquisition

Using a g.Nautilus Wireless Research EEG headset, EEG was recorded from 16 channels with active electrodes [27]. The reference electrode was fixed on the right earlobe and the ground electrode was positioned over the AFz electrode location according to the international 10/20 EEG standard. The EEG was filtered (Butterworth, 0.5-100Hz, eighth order), and sampled (sampling rate: 250Hz, down-sampled to 125Hz).

The BCI 2000 system was used to present the ‘oddball’ paradigm and to manage EEG data acquisition the graphical user interface (GUI) allowed real-time monitoring of the EEG signals and the management of stimulus trigger inputs. The g.TRIGbox [28] generated the trigger pulses for each of the auditory input signals. The g.Nautilus base station received the digitised EEG signals via wireless communication and connected to the g.TRIGbox via a VGA cable to integrate the EEG and stimulus trigger information. A splitter cable sent the synchronised trigger signals to both the BCI 2000 software and the headphones (delivering the audio to participants) – to accurately align the recorded EEG signals with the stimulus triggers.

EEG data acquisition and online signal processing, for the motor-imagery protocol, involved communication between a MATLAB Simulink [29] module, and the experimental protocol controller application presented in the Unity 3D Game Engine [30] – which was managed using a user datagram protocol (UDP) based communication.

### 2.4 Auditory oddball data analysis

#### EEG data preprocessing

Custom software, and EEGLAB [31] in Matlab, were used to analyse the auditory oddball recorded EEG data. The EEG signal was first high-pass filtered at 1Hz, secondly the 50Hz line noise was removed, thirdly anti-alias filtering was applied, and lastly, the cleaned signal was downsampled to 200Hz. Further preprocessing involved removing transient high-amplitude artifacts from the continuous EEG signal using the artifact subspace reconstruction method (ASR) [32], and performing an independent component analysis (ICA) to remove smaller artifacts related to eye-blinks, cardiac activity and muscle contractions, using the infomax algorithm [33].

#### Auditory oddball event-related potentials (ERPs)

Trials were time-locked to the stimulus onset and epoched between 100ms pre-stimulus onset and 1000ms post-stimulus onset. Epoched trials were baseline corrected, i.e., the mean voltage for the 100ms pre-stimulus interval was subtracted, low-pass filtered at 20Hz, and then averaged. The subsequent trial data derived from the vertex electrode (Cz), was used to extract the N1, N2, and P3 ERP component data. For the N1 component analysis, only the standard trials were used. The signal-to noise ratio (SNR) was computed by dividing the mean voltage signal by its own trial-to-trial standard at each time sample, to correct the effect of noise. Each participant’s N1 was thus computed as the largest negative SNR value (normalised using z scores) at the Cz electrode location, in the 80 – 180 ms post-stimulus time window. The post-stimulus time windows used or the N2 and P3 components analyses, were 180-400 ms and 200-450 ms, respectively. For both components, the average response to deviant/novel stimuli minus the average response to standard tones provided a measure of the magnitude of the oddball difference waveforms – and was calculated as the variance in the largest negative SNR for the N2 and the largest positive SNR for the P3, at the Cz electrode location and normalised using the z-scores. The standard error of the difference wave was computed as the sum of the trial-to-trial variances of the deviant trial amplitudes and standard trial amplitudes, i.e., the variance of differences. For all three ERP components, the latency of the peak amplitudes were also derived. The time windows selected for the N1 and N2 components differs from those implemented in the Kim et al. (2022) analysis – which were specific to the longer latencies of auditory evoked potentials in a paediatric population [23,34]. In relation to the auditory evoked N1, N2 and P3 components, the normal adult latencies ranges are approximately 80 – 120 ms, 160 – 270 ms, and 220 – 360 ms, respectively [34–36]. As the cohort for the current study were adults, who had varying levels of conscious awareness, the time windows were started at the approximate minimum post-stimulus latency for each component, but extended beyond the normal range, to allow for a delayed response due to brain injury – note, the minimum selected for the N2 time window was 20ms later than the approximate normal minimum, to allow for an extended N1 time window that did not overlap with the N2 minimum used.

### 2.5 Motor-Imagery Brain-Computer Interface (BCI) data analysis

A brief overview of the signal processing methods implemented in the MI-BCI framework is provided here, and full details are available in Coyle et al. (2022, preprint) [20].

#### Offline Signal Processing

EEG signals were processed using a filter-bank common spatial patterns (FBCSP) [37] and mutual information (MI) feature selection framework [38]. Signals were band-pass filtered into six frequency bands, and epochs were extracted for task intervals. CSP filters were calibrated to maximise class discriminability, and features were extracted using log-variance with a sliding window.

The regularised linear discriminant analysis (RLDA, from the RCSP toolbox[39]) classifier was then used to classify imagined movement and decoding accuracy (DA) was estimated across trials and runs. The peak DA was identified within specified intervals by selecting the highest local maximum within a 300ms window centred at the smoothed-reference and smoothed-task DA peaks. This approach ensured that the peak DA was not a random spike.

#### Offline Single-Run Analysis for BCI Calibration

Six-fold cross-validation was used to select the optimal channel set, frequency band set, classification window width, and feature number for each participant and run. DA was calculated and plotted as time-varying DA. The significance and robustness of DA were evaluated using permutation tests – which involved randomising the class labels and repeating the cross-validation process 100 times, creating a distribution of DA scores under the null hypothesis. The actual DA scores were then compared to this distribution to determine if they were significantly higher than what would be expected by chance with a 95% confidence interval.

Additionally, to provide further evidence that the peaks accuracy is significantly greater than chance (i.e., the DA for the run is significant) we also compare the mean peak DA value during the event-related task period to the baseline period using a dependent one-tailed *t*-test. Combining measures to determine significant runs ensures a reliable and robust measure of individual classification performance and clear evidence of engagement in MI by the participant or lack thereof (in the case of runs observed not be significant).

#### Online BCI Configuration

The calibrated FBCSP-MI framework’s optimised parameters were used in online runs to provide real-time auditory feedback. The classifier output, referred to as the time-varying signed distance (TSD), determined the movement direction and confidence of the audio feedback. TSD values were de-biased for stability, providing consistent feedback during the BCI task.

### 2.6 Statistical analysis

Statistical tests were performed using the IBM Statistical Package for the Social Sciences version 27.0 (SPSS). The dependant variables for each measure were:

- N1 component amplitude; measured for each participant as the largest negative SNR value (normalised using z-scores) at the Cz electrode location, in the 80–180 ms post-stimulus time window, averaged across standard stimulus trials in each run.
- N1 component latency; measured for each participant as the time (in milliseconds) of the largest negative value of the SNR at electrode location Cz in the 80–180 ms post-stimulus time window, averaged across standard stimulus trials in each run.
- N2 and P3 component magnitude; measured for each participant as the difference wave variance in the largest negative SNR for the N2 in the 180–200 ms post-stimulus time window and the largest positive SNR for the P3 in the 200–450 ms post-stimulus time window, at the Cz electrode location and normalised using z-scores, for trials in each run.
- N2 and P3 component latencies; measured for each participant as the time (in milliseconds) of the largest negative value of the SNR for the N2 in the 180–200 ms post-stimulus onset interval, and the largest positive value of the SNR for the P3 in the 200–450 ms post-stimulus onset interval, at electrode location Cz, averaged for each run.
- MI decoding accuracy (DA); measured for each participant as the percentage (%) of correctly classified trials out of the total number of trials in a run. Only DA values obtained from runs with a significantly higher mean DA peak (one-tailed t-test, *p* < 0.05) than maximum DA during reference baseline interval as well as DA achieved in randomly permutated trials were included in analyses. These data are referred to as significant runs.

The data were checked to determine whether analysis of variance (ANOVA) assumptions were met. For each analysis, the data were split according to group (UWS, MCS, and LIS) and the Shapiro-Wilk test was performed to check the normality of the data distributions. When found to be non-normal (Shapiro-Wilk (*W*), *p* < 0.05) for one or more groups, and/or skewness and kurtosis z-scores were found to be outside the accepted range (−1.96 ≥ 0 ≤ 1.96), or when outliers were present, a Kruskal-Wallis H nonparametric test was used. A Welch Test was chosen for analysis of data with a normal distribution when the variance was not homogenous (Levene’s statistic, *p* < 0.05), and pairwise comparisons were performed using a Games’ Howell test. When all ANOVA assumptions were met, a one-way ANOVA was performed, and the Tukey test was used to examine pairwise comparisons. Statistical tests of variance were two-tailed. The appropriate effect size calculations were performed for significant statistics; eta squared (*Ƞ^2^*) for ANOVA *F*-statistics, epsilon squared (*ε²*) for Kruskal-Wallis *H*-statistics and Omega Squared (ω^2^) for the Welch *W*-statistic. The range for each of these effect size measures is very small (> 0.01), small (0.01-0.05), moderate (0.06-0.13), and large (> 0.14) [40]. Using this approach, a one-way ANOVA was used to examine group differences in N1 and P3 latency as well as P3 difference wave magnitude, and Welch ANOVA was applied to examine group differences in N1 amplitude and N2 difference wave magnitude, and a Kruskal-Wallis test was used to examine group differences in N2 latency. The latter test was chosen due to outliers – one in the UWS data, and two in the MCS data. Given the small sample size, and likelihood of the outlier latencies being meaningful values, they were not removed. A two-tailed Spearman’s Rank Correlation test was chosen for bivariate correlation analyses, also due to the small sample size, and to mitigate the influence of a non-normal data distribution.

## 3 Results

### 3.1 Individual protocols

The main results are detailed in Supplementary Table 1 and are visually represented in Figure 2.

**Figure 1.**
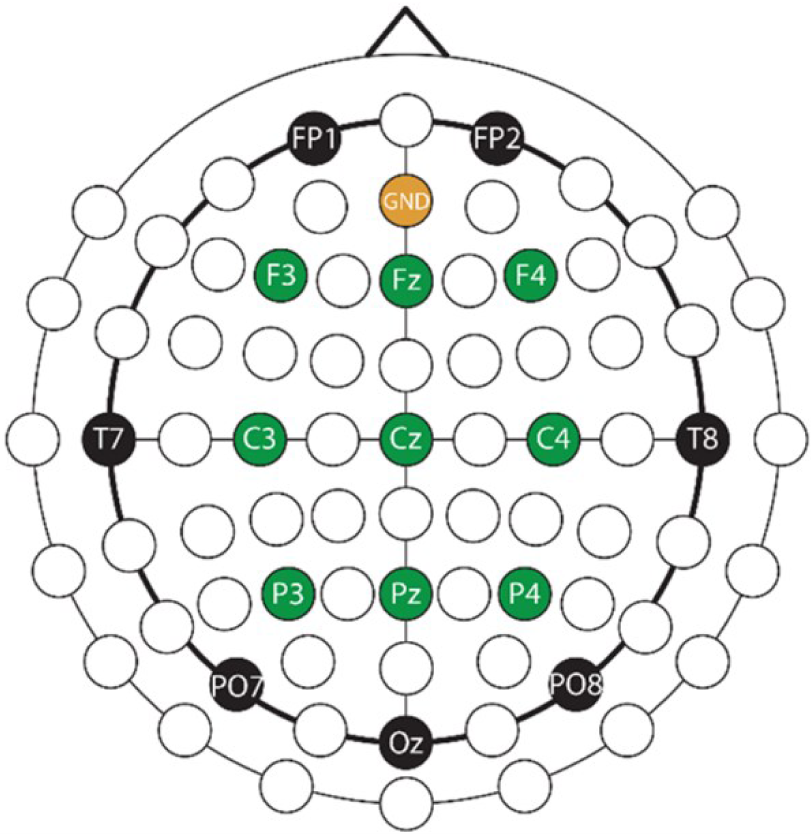
EEG montage. Illustration of the EEG and ground electrodes positions. Nine EEG channels covering motor and imagined movement-related cortical areas are indicated in green.

**Figure 2.**
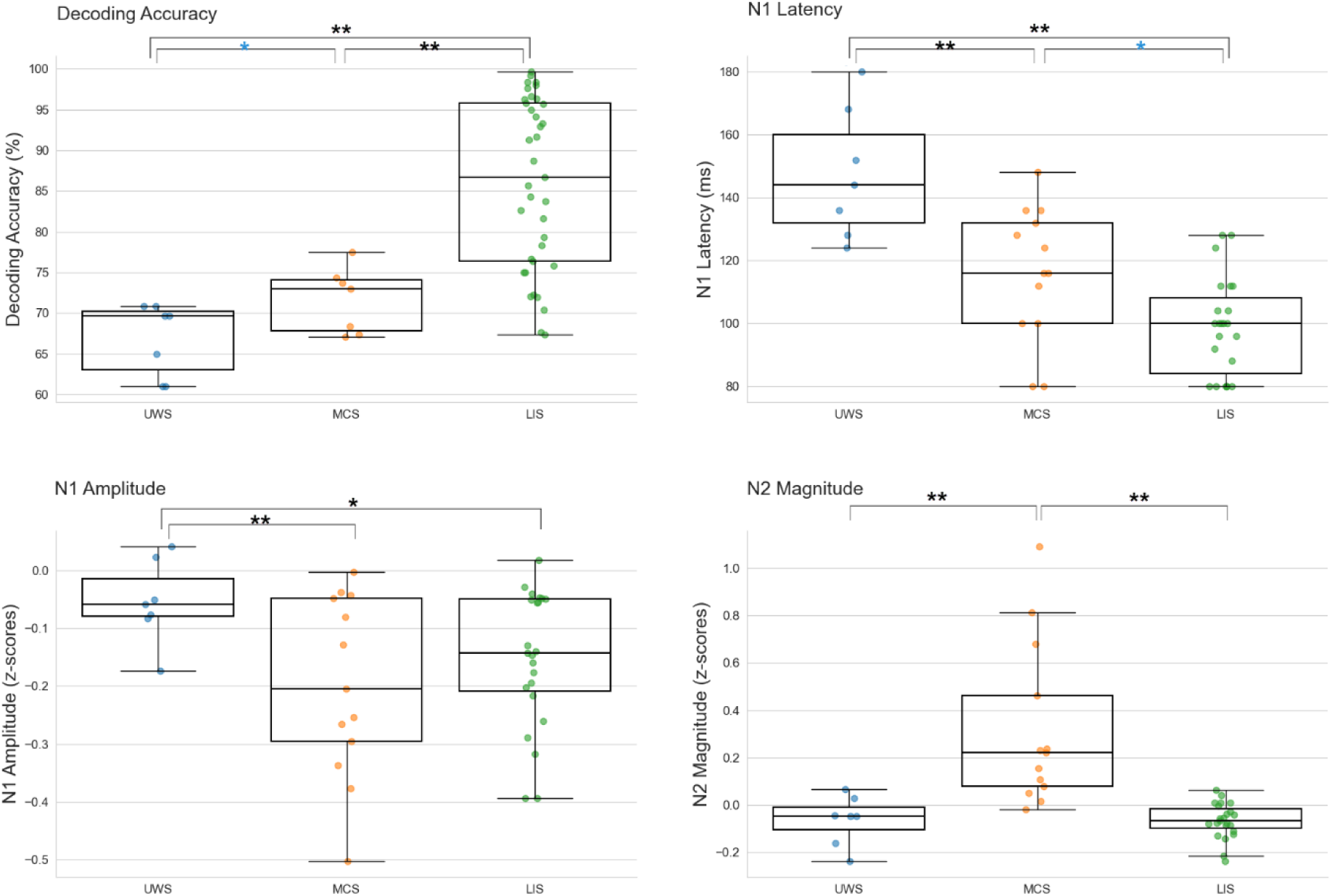
Illustration of the significant results. The dependent variable measure is presented on the y-axis, and group is labelled on the x-axis. Top left – Decoding Accuracy (DA, %); Significant group differences in MI-BCI decoding accuracies for significant runs. Top right – N1 latency (ms); Significant group differences in N1 latencies. Bottom left – N1 amplitude; Significant group differences in N1 amplitudes (z-scores). Bottom right – N2 magnitude; Significant group differences in the N2 difference wave magnitudes (z-scores). The whiskers extend to 1.5 times the interquartile range (IQR). A single black asterisk signifies a *p* value < 0.05, and a double black asterisk signifies a *p* value < 0.01, while the single blue asterisk indicates the difference approached significance at an alpha threshold of 0.05.

#### MI-BCI

Analysis of the DA scores, from significant runs within corresponding sessions, revealed a significant group difference (*W*_(2, 20.8)_ = 36.817, *p* < 0.001, *ω^2^* = 0.743). According to a Games-Howel comparison, the significant result was a consequence of the higher MI performance of the LIS group compared to both the MCS (*MD* = 13.99, *SE* = 2.15, *p* < 0.001) and UWS (*MD* = 19.15, *SE* = 2.25, *p* < 0.001) groups, while the higher MCS performance compared to the UWS performance, approached significance (*MD* = 5.156, *SE* = 1.99, *p* = 0.053).

#### Oddball Paradigm

Group differences in the amplitude of the N1 ERP were found to be significant (*F*_(2, 21.246)_ = 7.041, *p* = 0.005, *ω^2^* = 0.324), due to a significantly lower amplitude for the UWS group compared to the MCS group (*MD* = 0.176, *SE* = 0.052, *p* = 0.007), and the LIS group (*MD* = 0.109, *SE* = 0.037, *p* = 0.022). The latency of the N1 was also significantly different across groups (*F*_(2, 43)_ = 20.42, *p* < 0.001, *Ƞ^2^* = 0.487), with UWS N1 latency found to be significantly longer compared to the MCS (*MD* = 33.07, *SE* = 8.18, *p* < 0.001), and LIS (*MD* = 48, *SE* = 7.53, *p* < 0.001) groups. Additionally, the difference between the MCS and LIS groups was just above the alpha threshold (*p* = 0.053, longer for MCS).

The magnitude of the N2 response was also found to differ significantly across groups. A Welch ANOVA revealed the group difference in the magnitude of the N2 ERP (*F*_(2, 15.02)_ = 8.304, *p* = 0.004, *ω^2^*= 0.434) was due to a significantly larger mean difference wave magnitude for the MCS group compared to both the UWS ( *MD* = 0.438, *SE* = 0.11, *p* = 0.003) and LIS (*MD* = 0.441, *SE* = 0.106, *p* = 0.003) groups. The group difference in N2 latency was not found to be significant (*H* = 3.035, *p* = 0.219).

A significant group difference was not found for either the magnitude of the P3 difference wave (*F*_(2, 43)_ = 0.431, *p* = 0.653) or the latency of the P3 component (*F*_(2, 43)_ = 1.385, *p* = 0.261).

### 3.2 Relationship between protocol metrics

To assess whether there was convergence between the findings of the significant ERP component group differences and the DA group differences, we conducted a two-tailed Spearman’s Rank bivariate correlation analysis to examine the relationship between the DA scores for significant runs and each of the following variables; N1 amplitude and latency, and N2 difference wave magnitude. The results show the DA scores are significantly negatively correlated with N1 latencies only (*ρ* = −0.802, *p* = 0.017, see Figure 3).

**Figure 3.**
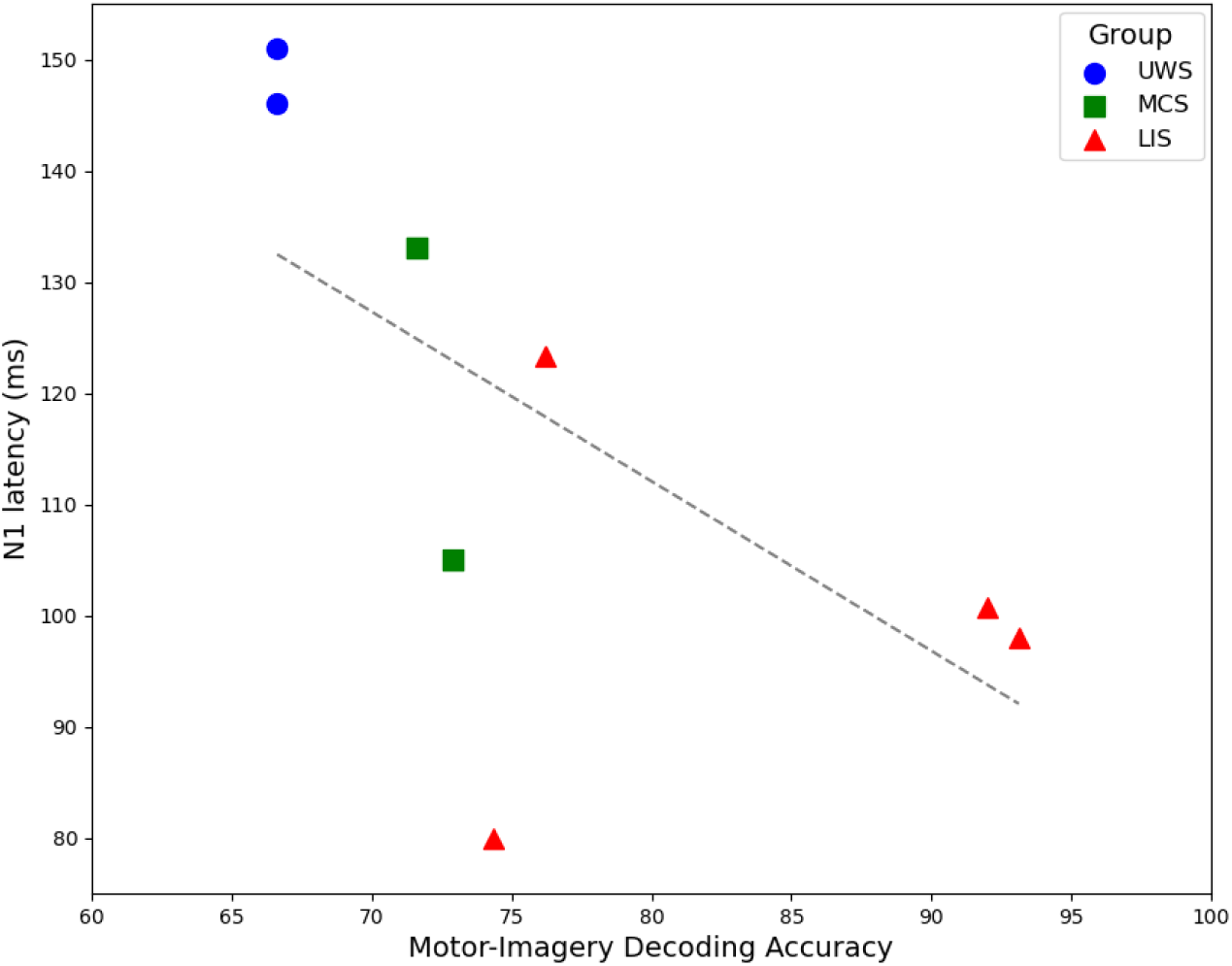
Illustration of the significant negative correlation between individual mean N1 latencies and corresponding mean DA scores for significant runs, i.e., the peak DA during the task period was significantly higher than the baseline period – across the UWS (blue circles), MCS (green squares), and LIS (red triangles) groups.

**Figure 4.**
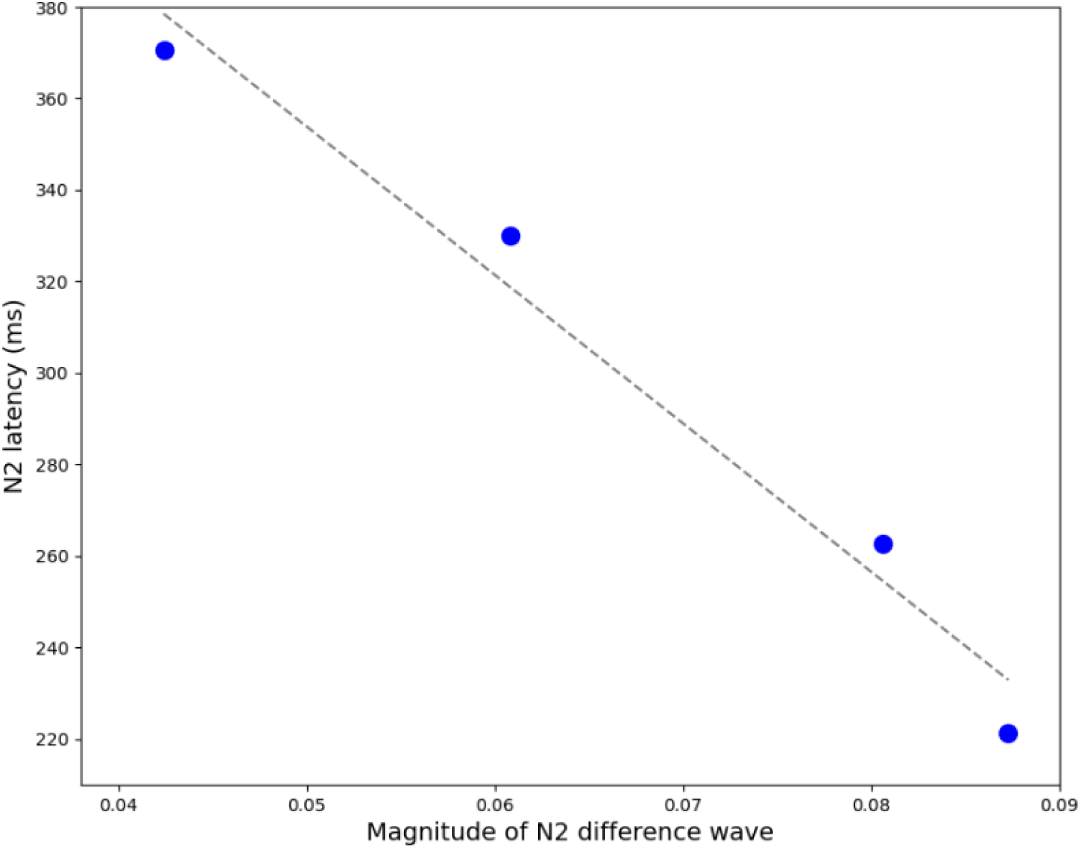
LIS patients only. Illustration of the relationship between the N2 difference wave magnitude (x-axis; calculated as averaged z-scores for each individual), and N2 latency (y-axis; calculated as the average peak latency for ERP’s time-locked to stimulus presentation, 180-400ms post-stimulus onset).

**Figure 5.**
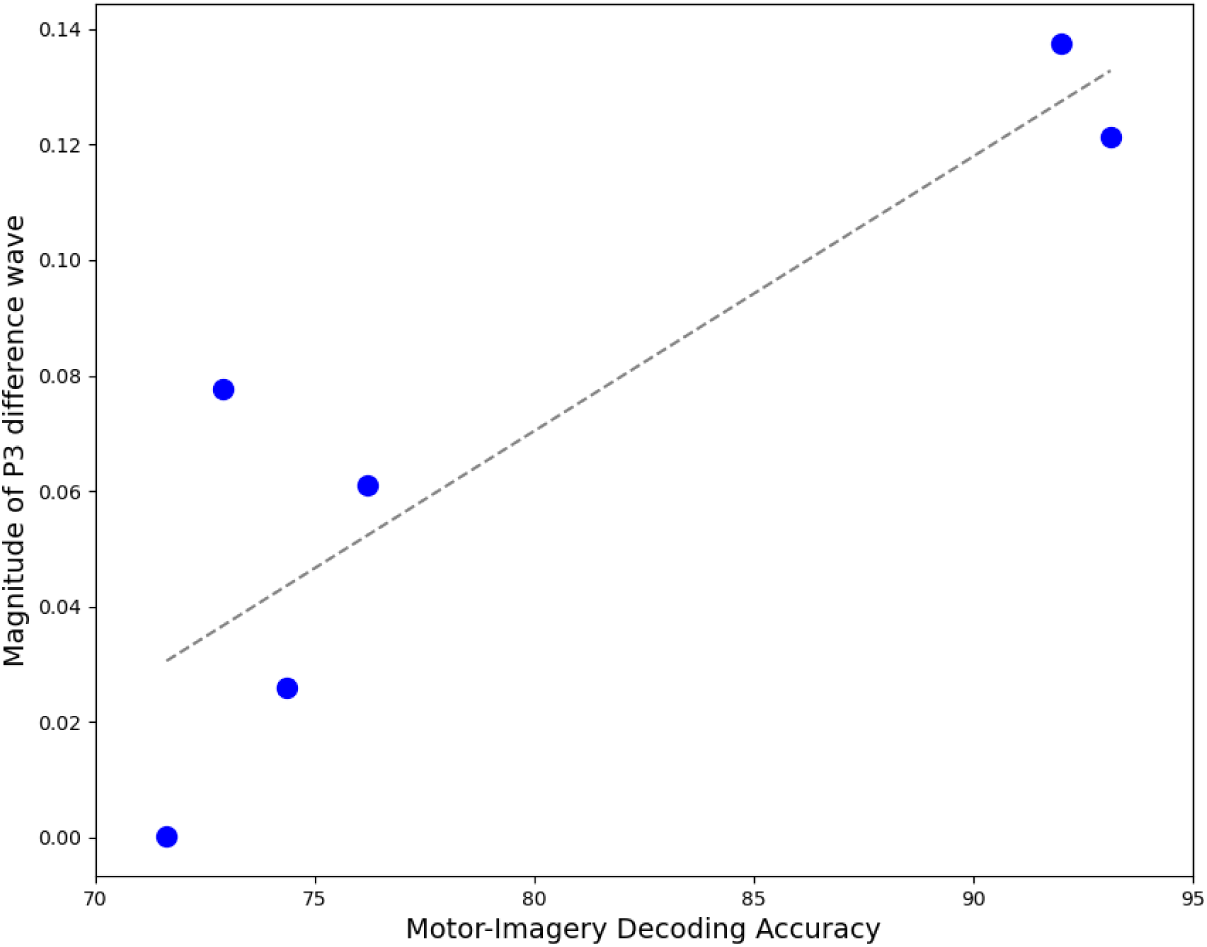
MCS and LIS patients only. Illustration of the relationship between the magnitude of the P3 difference wave (y-axis; calculated as averaged z-scores for each individual), and corresponding DA scores (x-axis; calculated as averaged DA for significant MI trials).

**Figure 6.**
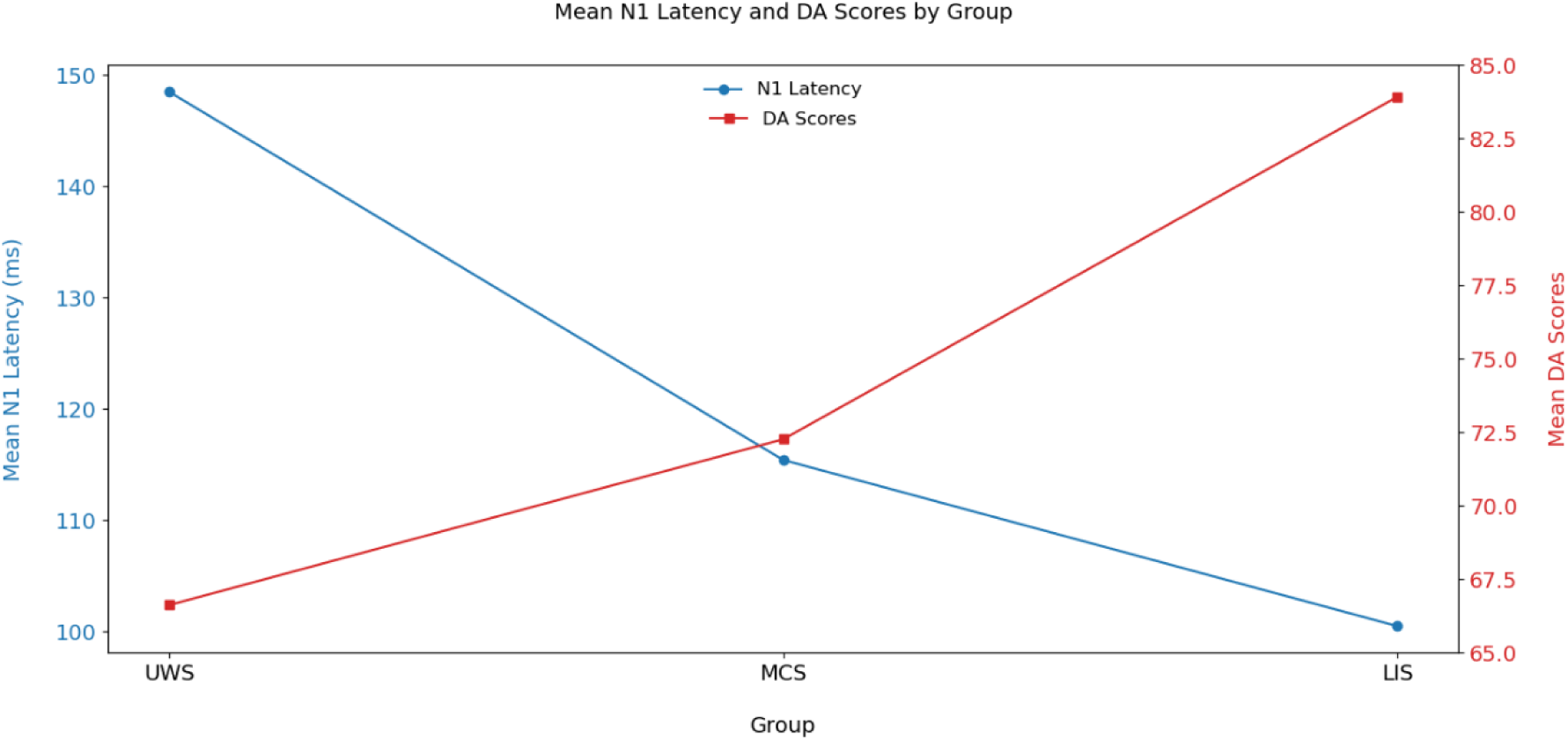
Illustration of the group means for N1 latency (ms) in blue (with circle markers) and group mean scores for decoding accuracy (DA) in red (with square markers) on trials that were significant, i.e., the peak DA during the task period was significantly higher than the baseline period.

The findings from the combined oddball and MI-BCI protocol, implemented over two to three sessions with a pilot sample of five PDoC and four LIS patients, highlight significant group differences in the latency and strength of the auditory evoked N1 ERP. The results indicate that, similar to its effectiveness in distinguishing between cognitive states in brain-injured paediatric groups [23], the amplitude of the auditory evoked N1 ERP component can serve as a potential EEG marker to aid in diagnosing UWS in adult PDoC patients (*p* = 0.005). However, the N1 amplitude did not clearly separate MCS and LIS groups (LIS v MCS; *MD* = 0.067, *SE* = 0.049, *p* = 0.379), and while the N2 difference wave magnitude was found to differentiate between MCS and UWS (*p* = 0.005), and MCS and LIS (*p* = 0.003) groups, this magnitude measure was remarkably similar for LIS and UWS patients (*p* = 0.648). Further differing from the paediatric group of brain injured patients in the Kim et al. (2022) study [23], the adult PDoC patient groups in the current study did not differ significantly according to the P3 difference wave magnitude, or in P3 latency, across groups.

Underlying the significant N1 ERP latency result for this cohort of PDoC and LIS patients, is an observable linear increase in the latency of the N1 ERP responses, time-locked to the stimulus 100.5ms (±16.9) post-onset for the LIS group, and increases linearly to 115.43ms (±20.5) for MCS patients and 148.5ms (±19.4) for UWS patients (see **Error! Reference source not found.**). Taken as a whole, the evaluation of group differences that relate to the N2 component indicate a significantly greater N2 difference wave magnitude for the MCS patient group, with a latency within the normal range (220 – 270 ms [34]). The reduced N2 magnitude and later latency, found for the UWS group was anticipated.

Counterintuitively, the LIS group N2 difference wave magnitude differed only fractionally from that of the UWS group, and with increased latency (group mean; 296.17ms). However, it is noted that the longer latency was influenced by two of the four LIS group participants (average individual N2 latencies were; 262.67, 370.67, 330, 221.33). When the N2 difference wave magnitude and N2 latency data for the LIS group are graphed to illustrate their relationship, the resulting plot demonstrates an almost perfect linear correlation (see **Error! Reference source not found.**). For illustration purposes, the sign o f the z scores (N2 magnitude values) was changed to positive, to highlight the increase in magnitude as the N2 latency is closer to normal (∼200ms). While this is an interesting observation and warrants further investigation with a larger sample, the current analysis indicates that the N2 ERP component does not effectively distinguish between UWS, MCS, and LIS patients.

Regarding the P3 ERP results, Kim et al. (2022) [23], found, within a paediatric cohort with brain injury, that the magnitude of the P3 difference wave was positively correlated with the individual’s Motor Command-Following (MCF) ability, as measured by significant power spectral differences between rest and movement conditions in the alpha-theta frequency bandwidth. However, the results presented here do not find significant group differences in the magnitude of the P3 difference wave, or the latency of the component – and a Spearman’s Rank correlation test did not find a significant correlation between either P3 component measure and corresponding DA for significant MI trials.

However, given the length of the window used to calculate the P3 latency (200 – 450 ms), the UWS mean group P3 latency of 260ms may reflect a delayed P2 ERP component of the N1/P2 complex, which has a normal latency range of 50-120 ms [41] – a response associated with changes in the auditory environment, such as deviations in the frequency or intensity of a sound [41,42]. When confined to LIS and MCS patients only, a significant one-tailed positive correlation is found between the magnitude of the P3 difference wave and the DA scores for significant runs (*ρ* = 0.771, *p* = 0.036, note; a one-tailed Spearman’s Rank correlation was justified based on the hypothesis that the correlation would be positive, as deduced from the Kim et al., 2022, study results [23]). This result suggests that the magnitude of a P3 difference wave, when the P3 ERP component exhibits a normal post-stimulus latency (∼300ms), could provide an objective marker of an individual’s capacity to perform motor-imagery – further research with a larger participant cohort is required to test this hypothesis.

Overall, within this cohort, the N1 latency was identified as the EEG-marker that most distinctly differed across groups – with significantly longer latencies for UWS compared to MCS (*p* < 0.001) and LIS (*p* < 0.001) groups, and longer latencies for MCS compared to LIS patients, with a difference that approached significance (*p* = 0.053). Similarly, regarding, differences in the ability to modulate sensorimotor rhythms with imagined movements, the DA for significant MI runs was found to effectively separate each group, with higher DA for the LIS group compared to the MCS group (*p* < 0.001) and the UWS group (*p* < 0.001), and with greater DA for the MCS group compared to the UWS group, with the difference tending towards significance (*p* = 0.053).

As can be seen in **Error! Reference source not found.**, the trend between MI-BCI DA and group assignment and the trend between N1 ERP latency and group assignment, are anti-correlated, i.e., lower DA is linked to longer N1 latency and higher DA is linked to shorter N1 latency and a significant negative correlation between N1 latency and significant DA scores (*p* = 0.017) is observed. Furthermore, group differences in DA and the N1 latency, were found to have large effect sizes (0.743 and 0.487, respectively).

## 4 Discussion

Currently, misdiagnosis rates are extremely high for patients with a DoC – reportedly, 43% of MCS patients are diagnosed as UWS) [43]. Fins et al. (2020) have noted a discrepancy between the advancements in scientific understanding of DoC and actual clinical practice, highlighting the need for better diagnostic tools and therapeutic interventions to be more widely implemented in clinical settings [44]. Higher cognitive functions depend on the integrity of large-scale intrinsic cortical networks (ICNs) [45], and the overall functional status and level of consciousness in patients with diffuse brain injuries has been correlated with electrophysiological measures [46]. Thus, advanced neuroimaging techniques and electrophysiological biomarkers offer the most promise in addressing this discrepancy [45,47,48]. EEG stands out as the most cost effective neuroimaging modality, and recent advances in wearable EEG systems allow bedside applications, increasing accessibility – while the range of EEG-based methods demonstrated to provide diagnostic value further promotes the utility of EEG-based assessments for this cohort [22,45,49]. However, the importance of combining various EEG methods to offer a comprehensive assessment of a patient’s brain function to aid in more accurate diagnoses and prognoses have recently become more central to research in this area [49,50]. Leading methods include evaluation of; ERP components, EEG modulation associated with motor-imagery tasks, and frontoparietal alpha network connectivity [48,49,51]. A further consideration related to the delivery of care within this cohort, is their inability to communicate, which has a significant impact on wellbeing and excludes the patient from the decision-making around their care highlighting the importance of BCIs to address this deficit [44,52].

In line with this research focus, the pilot study presented here has evaluated the potential for the combined MI-BCI and passive auditory oddball EEG-based protocols to enhance the diagnostic accuracy for PDoC states and LIS. The findings demonstrate high internal consistency between the N1 latency and MI DA scores (*ρ* = −0.802), corroborating the reliability and effectiveness of these neurophysiological markers – offering robust evidence for their use as objective, movement-independent measures of consciousness. While establishing internal consistency is an important step in the validation process for a neuroimaging toolkit to aid in the diagnosis of PDoC state – this validation relies on a group-based variation, which does not account for individual heterogeneity. Accurate assessment tools need to have good sensitivity (i.e., a low rate of false negatives) and specificity (i.e., a low rate of false positives) [50], and therefore, there is a need to accommodate individualised pathology. A recent gap-analysis concludes that the heterogeneity in DoC pathology requires domain-specific brain function analyses rather than generalised behavioural checklists, as these targeted analyses are more likely to identify subsets of patients with covert consciousness who can wilfully modulate brain activity, despite being unable to demonstrate overt expressions of awareness – thereby establishing ‘BCI readiness’ [53]. Furthermore, Schiff et al. (2024), emphasise the importance of personalised BCI solutions for this subgroup of patients once they are no longer in the acute phase, that are tailored to home environments and patient preferences [53]. The current MI-BCI protocol includes an assessment phase to evaluate the individual’s capacity to wilfully modulate their brain activity, determined by achieving a significant DA in at least one of two assessment runs – thus establishing the patient engagement and readiness. Importantly, the MI analysis reported here covers as little as two or three sessions – an early (third or fourth) session and one or two later sessions (following eight to ten training/feedback or Q&A sessions). Although the later sessions followed several others, given there are 60 trials in a training or feedback run and 48 in a Q&A run, with four runs per session, the findings indicate that three sessions with this structure provides sufficient data to achieve separability of PDoC states. While the protocol offers a range of music genres as auditory feedback, there is scope to further personalise the feedback for individual users. Furthermore, the addition of data on the latency of individual’s N1 ERP component, provided by the passive auditory oddball task, improves the reliability of a potential diagnosis.

Guided by research-informed recommendations, and the results reported here, future research should continue to expand the neuroimaging toolkit to provide a comprehensive assessment based on ERP signatures and frontoparietal alpha network connectivity that demonstrate strong internal consistency. The development of such a toolkit will aid in the diagnosis of DoC states, with a reduced likelihood of misdiagnosis due the loss of overt behaviour or to variations in individual pathology that do not match the group phenotype – improving sensitivity and specificity. Regarding prognostic assessment, given the sample included in this pilot study were all more than 4-weeks post injury and were therefore in the prolonged stage of DoC, future longitudinal research, should consider evaluating the relationship between MI ability and N1 latency in the early stages of DoC, and at specific timepoints through the first year of recovery. This prospective research would establish the potential prognostic value of the N1 latency – which would provide further support for the use case of this neuroimaging protocol within this cohort [9]. Further investigation of N2 and P3 ERP component parameters in relation to LIS, and LIS plus MCS patients, respectively, is also warranted with a larger patient cohort.

### 4.1 Limitations

The findings of this pilot study were underpowered, with the main results for group differences in DA for significant MI trials, N1 latency, and the correlation between these variables, achieving 32%, 16%, and 37% power, respectively – based on a post-hoc analysis in G*power [54]. However, based on the effect sizes for these results (DA: ***ω^2^*** = 0.743; N1 latency: *Ƞ^2^*= 0.487; significant DA and N1 latency correlation: *ρ =* −0.802) – a minimum-maximum total sample size to achieve 80% power is 21-45 – according to an a priori power analysis in G*power. A further limitation stems from the presentation of the auditory oddball protocol. There were five different runs to choose from, each with different novel sounds, mixed with deviants and standards in a fixed random order. In a given session, two runs were selected, and each run was delivered for a duration of five-minutes, in a loop. This resulted in four repetitions of a given run – which could have influenced the magnitude of the later N2 and P3 auditory ERP components, as the stimuli were not completely randomised, resulting in a pattern that could potentially have been learned. The N2-P3 complex is associated with change-detection – the N2 component is evoked pre-attentively, by changes in the physical features of a stimulus, and is referred to as the mismatch negativity (MMN) response, while the P3 (particularly the P3a) ERP component that follows MMN, reflects the reorientation of attention to a deviant stimulus [55]. However, the N2 and P3 stimulus evoked responses have been found to be robustly automatic, and therefore resilient to increased frequency and/or regularity of deviant stimuli [56].

Despite the limitations of this pilot study, the significant results support further investigation within a large cohort of PDoC and LIS patients, to determine the diagnostic and prognostic potential for these objective, movement-independent EEG-markers, measurable using the MI-BCI and passive auditory oddball protocols described here.

## 5 Conclusion

The results of this pilot study provide evidence to support the applicability of the EEG-markers identified by Kim et al (2022) [23], within a cohort of adult PDoC patients, as objective neurophysiological, and movement independent measures that can aid the assessment of conscious state within a cohort of adult PDoC and LIS patients. While a reduced N1 amplitude has been indicated as a potential EEG-marker for UWS, the N1 latency emerges as a more robust metric for adult PDoC patients – exhibiting a longer mean latency for UWS compared to both MCS and LIS groups, that is significant at an alpha level of 0.001, and has a large effect size (*Ƞ^2^* = 0.487). Furthermore, the difference between the MCS and LIS groups mean N1 latency approached significance (*p* = 0.053). Furthermore, the mean DA for significant MI trials was demonstrated to similarly separate groups, again reinforced by a large effect size (*ω^2^* = 0.743).

Importantly, the N1 latency was found to have a strong negative correlation with DA scores (*ρ* = −0.802, *p* = 0.017), with shorter latencies (closer to normal) linked to better DA – indicating high internal consistency between the N1 latency and the MI DA scores, which corroborates the potential effectiveness of these combined protocols in providing robust and objective measures of awareness in PDoC. Thus, the findings presented here strongly support the combined use of these MI-BCI and passive auditory oddball protocols, as a tool to aid in diagnosing PDoC states and LIS.

## Data Availability

The data supporting the findings of this study are available from the corresponding author upon reasonable request.

## Supplementary Material

**Supplementary Table 1.** Oddball. Peak amplitude of the N1 ERP, and magnitude of the N2 and P3, and Latency of all three ERP components—Mean (SD) and statistics, across groups. The reported statistics are the F statistic for one-way ANOVA and Welch ANOVA analyses, and the H statistic for the Kruskal-Wallis analyses. Effect sizes are reported as Eta Squared (*Ƞ^2^*) for ANOVA statistics, Omega Squared for Welch statistics (*ω^2^*), and epsilon squared (*ε²*) for H statistics.

|  |  | UWS |  | MCS |  | LIS |  | Statistics |  |  |
| --- | --- | --- | --- | --- | --- | --- | --- | --- | --- | --- |
|  |  | *n* = 2 |  | *n* = 3 |  | *n* = 4 |  |  |  |  |
| EEG | Measure | *M* | *SD* | *M* | *SD* | *M* | *SD* | (*stat*)<br>val | *p* val | $\eta^2/ \varepsilon^2/ \omega^2$ |
| N1 | Amplitude (1zB) | -0.039 | 0.079 | -0.215 | 0.162 | -0.147 | 0.117 | (*F*)<br>7.041 | 0.005 | 0.324 |
|  | Latency (ms) | 148.5 | 19.413 | 115.43 | 20.508 | 100.5 | 16.86 | (*F*)<br>20.42 | <0.001 | 0.487 |
| N2 | Magnitude (2-1zB) | -0.0642 | 0.0969 | 0.3733 | 0.3926 | -0.0678 | 0.0728 | (*F*)<br>8.304 | 0.004 | 0.434 |
|  | Latency (ms) | 327.5 | 79.468 | 258.29 | 74.449 | 296.17 | 89.384 | (*H*)<br>3.035 | 0.219 | – |
| P3 | Magnitude (2-1zB) | 0.064 | 0.0941 | 0.062 | 0.07 | 0.086 | 0.094 | (*F*)<br>0.431 | 0.653 | – |
|  | Latency (ms) | 260 | 74.466 | 327.14 | 95.861 | 314 | 97.972 | (*F*)<br>1.385 | 0.261 | – |
| MI | DA | 66.625 | 4.183 | 71.781 | 3.772 | 85.778 | 10.42 | (*F*)<br>36.82 | <0.001 | 0.743 |

1zB; the z scores for the amplitude of the standard stimuli, baseline corrected
2-1zB; the z score for the difference wave, baseline corrected
DA; Decoding Accuracies

**Supplementary Table 2.**
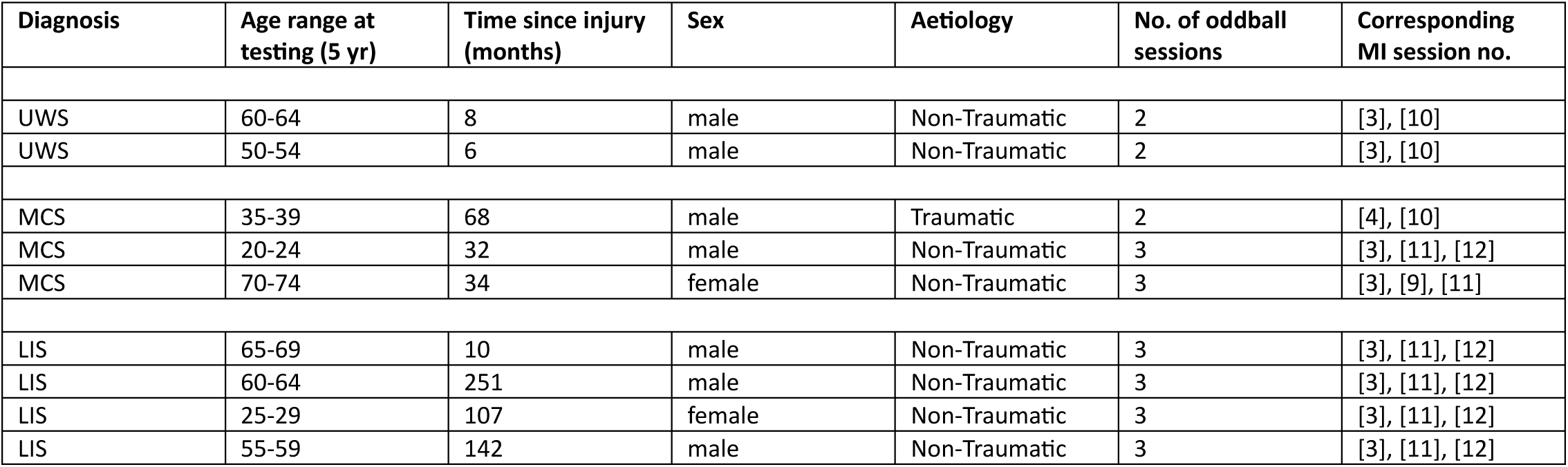
Participant and session details.

